# Comprehensive analysis of Histone deacetylases genes in the prognosis and immune infiltration of glioma patients

**DOI:** 10.1101/2022.01.24.22269795

**Authors:** Aibin Liu, Yanyan Li, Lin Shen, Na Li, Liangfang Shen, Zhanzhan Li

## Abstract

The occurrence and development of tumors are closely related to histone deacetylases (HDACs). However, the overall biology and prognosis are still unknown in glioma. In the present study, we comprehensively explored the biology function and prognosis of eleven HDAC genes in glioma, which may contribute the more understanding of molecular mechanisms and potential therapeutic targets for glioma patients.We systematically described the expression files, molecular subtypes, prognostic value, immune filtration and tumor microenvironment and gene alteration, function and pathways enrichment, and drug sensitivity using TCGA and CGGA datasets. We developed and validated the prognostic model based on HDACs genes in glioma using LASSO, univariate, and multivariate cox regression. Receiver operating characteristic analyses were used for model evaluating. We also validated the expressions of HDACs genes included in the model in non-tumor and glioma tissues samples. Glioma patients can be divided into two subclasses based on eleven HDAC genes, and patients from two subclasses had markedly different survival outcomes. Then, using six HDAC genes (HDAC1, HDAC3, HDAC4, HDAC5, HDAC7, and HDAC9), we established a prognostic model in glioma patients, and this prognostic model was well validated in an independent cohort population. Furthermore, the calculated risk score from six HDACA genes expression was suggested to be an independent prognostic factor, which can predict the five-year overall survival of glioma patients well. High-risk patients can be attributed to multiple complex function and molecular signaling pathways, and the genes alterations of high- and low-risk patients were significantly different. We also found that different survival outcomes of high- and low-risk patients could be involved in the differences of immune filtration level and tumor microenvironment. Subsequently, we identified several small molecular compounds that could be favorable for glioma patients’ treatment. And finally, the expression levels of HDAC genes from prognostic model were validated in glioma and non-tumor tissues samples.Our results revealed the clinical utility and potential molecular mechanisms of HDAC genes in glioma. Model based on six HDAC genes can predict the overall survival of glioma patients well, which can be served as potential therapeutic targets.

## Introduction

Glioma was deemed to be the most aggressive tumor in central nervous system. The annual incidence of glioma was reported to be about 30-80/1 million in the world, and increased by 1-2% annually, the 5-year survival rate was only 10%-20%[1]. According to the WHO standard, glioma was divided into four grades by its pathological characteristics[2]: grade I, pilocytic astrocytoma, which manifested as benign tumor, patients may have full clinical recovery after total tumor resection. Grade II, which had a poor prognosis compared with grade I, but was still considered to be low-grade glioma. Grade III, such as anaplastic astrocytoma as well as Grade IV, GBM were types of advanced grade glioma lined to high degree of malignancy, strong invasive ability, poor prognosis and multiple differentiation potentials, the median survival time was only about one year[3]. Despite various cancer therapies have been applied over the past decades, the prognosis of glioma patients remains dismal. After combining the isocitrate dehydrogenase (IDH1/2) mutation and whether the 1p19q code is missing, the HWO classification of central nervous system tumors is more refined[4]. However, the clinical outcomes and side effects of patients with same grade and classification of tumors are not the same after being comprehensively treated[5]. This suggests that we still need to explore more instructive molecular targets in the research of glioma with the consideration of unclear and complex molecular mechanisms.

The underlying cause of malignant tumors is the disorder of gene expression system, including oncogenes, tumor suppressor genes and genes related to DNA repair[6]. With the continuous development of epigenetics, it is gradually recognized that almost all malignant tumors have epigenetic abnormalities, which together with gene changes cause tumorigenesis[7]. The epigenetic phenomena involved in the occurrence of malignant tumors mainly include abnormal DNA methylation, histone modification and their interaction caused by abnormal expression of non-coding RNA and chromosomal remodeling[8]. These epigenetic changes lead to abnormal activation of certain genes and silencing, thereby allowing cell growth to enter an uncontrolled state. The occurrence and development of tumors are closely related to histone deacetylases (HDACs)[9]. Studies have shown that genome-wide histone acetylation levels are generally reduced in tumor cells, among which HDAC1, 5 and 7 are regarded as tumor markers[10]. Second, studies have shown that gene knockout of HDAC1/2 in breast cancer cells or HDAC1/2/3 in colon cancer cells can induce tumor cell apoptosis, suggesting that the activity of HDACs is related to tumor cell survival[11, 12]. Similarly, abnormal binding of HDACs to oncogene fusion proteins to certain gene loci is also regarded as an important mechanism of tumorigenesis. The HDAC consists of four classes: class □ (HDAC1, HDAC2, HDAC3, HDAC8), class □ (HDAC4, HDAC5, HDAC6, HDAC7, HDAC9, HDAC10), class □ (SIRT1-SIRT7), and class □ (HDAC11)[13]. Previous study had explored the roles of HDACs in the prognosis of clear cell renal cell carcinoma[14]. However, the overall biology and prognosis are still unknown in glioma. In the present study, we comprehensively explored the biology function and prognosis of eleven HDAC genes in glioma, which may contribute the more understanding of molecular mechanisms and potential therapeutic targets for glioma patients.

## Materials and methods

### Data source

The mRNA expression data of glioma patients from The Cancer Genome Atlas (TCGA, https://portal.gdc.cancer.gov/) and Chinese Glioma Genome Atlas (CGGA, http://www.cgga.org.cn/). The gene mutation data were also from the TCGA database. Drug response data were available from the Genomics of Drug Sensitivity in Cancer (GDSC) database (https://www.cancerrxgene.org/downloads). The immune filtration data from TCIA (https://tcia.at/home). The histone deacetylases genes (HDAC1, HDAC2, HDAC2, HDAC4, HDAC5, HDAC6, HDAC7, HDAC8, HDAC9, HDAC10, HDAC11) were from the molecular signatures database (MSigDB).

### Clustering analysis

Using R “Consensus Cluster Plus” package, we performed the clustering analysis. The Consensus Cluster Plus allowed data clustering with negative value. Using K-means method, we achieved the most approximate number of clusters by extracting 1000 times from 80% of sample size. The results were presented using consensus matrix heatmap. We also used the principal component analysis (PCA) and the t-distributed stochastic neighbor embedding (tSNE) to further validate the clustering analysis.

### Development and validation of HDAC genes prognostic model

Using 11 HDAC genes, we developed an overall survival (OS) prognosis model in TCGA dataset. The Least Absolute Shrinkage and Selection Operator (LASSO) regression was used to select the number of entering the model, then a multivariate cox regression was performed to get the regression coefficient of each HDAC gene. We calculated the risk score of each sample using the following formula: risk score =coef_1_*gene1 expression+coef_2_* gene_2_ expression+…coef_n_*gene_n_ expression. Using the established prognostic model, we performed the validation in CGGA dataset. Glioma patients were separated into high- and low-risk group according the median of risk score. Kaplan-Meier survival cure to compare the difference of OS between high-and low-risk group. The receiver operating characteristic curve (ROC) was used to evaluate the predictive ability at 1-year, 2-year, and 3-year of patients’ OS of TCGA and CGGA. The PCA analysis was used to identify the risk type.

### Clinical correlation and independent and analysis

To investigate the association between risk score and prognosis in glioma patients, we first performed the stratified analysis in different clinical parameters, then we compared the difference of HDAC genes between two clustering groups, and the risk score distributions were also observed in different clinical parameters. To validate the independence of risk score, we performed univariate and multiple variate cox regression analyses by adjusting the clinic parameters (TCGA: age, gender, grade; CGGA: age, gender, grade, history, TNM stage, radiotherapy, chemotherapy, occurrence type, IDH and 1p19q status). We evaluated the diagnostic ability of risk score and other parameters using ROC. We built the nomograph to evaluate the clinical application of HDAC genes prognostic model, and the nomogram-predicted probability of 1-year OS, 3-year OS and 5-year OS were used to assess the model fitting ability.

### Functional, pathway enrichment and mutations analysis

To explore the function and pathway enrichment of different high- and low-risk groups, we performed the GO functional enrichment and KEEG pathway analysis using the “clusterProfiler” package. Using the masked copy number segmentation data, we investigate the gene mutations frequency of different risk group using the “maftool” package (gene alteration, variant classification, variant type, co-occurrence and mutually exclusive).

### Immune filtration, tumor microenvironment, and drug sensitivity analysis

We assessed the difference of 16 immune cells related infiltrating score and 13 immune-related pathways between high- and low-risk groups. Using TCGA dataset, we calculated the immune and stromal estimate scores of high- and low-risk groups using R “estimate” package. To explore the correlation between small molecular drugs and the identified prognostic signature genes, Pearson correlation coefficients were calculated. |R|>0.25 and P<0.05 were considered significantly correlated.

### Validation of HDAC genes in glioma and non-tumor tissue

To validate the expression of six HDAC genes included the prognostic model (HDAC1, HDAC3, HDAC4, HDAC5, HDAC7, and HDAC9), we detected the expression of these HDAC genes in glioma and non-tumor tissues. mRNA expression data were collected from 23 samples of epilepsy patients and 157 tumor samples. The tissue collection was approved by the NCI IRB committee with informed consent obtained from all subjects[15]. Quantitative polymerase chain reaction was performed to detect mRNA expression level of HDAC genes. These methods have been previously described. Briefly, Total RNA was isolated using TRIzol (Invitrogen). 1,000ng RNA was reverse-transcribed into cDNA in a total reaction volume of 20□l with PrimeScriptTM RT Reagent Kit according to manufacturer’s instructions (Takara). The qPCR primer sequences were designed by Santa-cruz Biotechnology Co, Inc. Real-time PCR was performed using SYBR premix Taq via CFX96 Real-Time PCR Detection System (Bio-Rad, Richmond, CA, USA). The 2-ΔΔCT method was used to calculate the relative expression of mRNA.

### Statistical analysis

Differentially expressed genes analysis were performed using “limma” package. Differences for category variables were performed using Chi-square test. The comparisons of OS curve were achieved using log-rank test. One-way ANOVA was used for comparing the differences of HDAC genes expression among non-tumor and different grade glioma. SNK methods was used for multiple comparisons. All statical analysis were finished using R software 4.0.1, and *P*<0.05 was considered significant.

## Results

### Identification of two subclasses in glioma

The flow chart was plotted to comprehensively describe our study (Figure 1A). For 11 HDAC genes, the correlations among HDAC members were different. HDAC6 and HDAC8 showed positive association with other HDAC genes, while HDAC3 and HDAC4 showed negative correlation with other HDAC genes (Figure 1B). We performed clustering analysis using 11 HDAC genes (HDAC1-HDAC11). The consistency coefficient was calculated to achieve the optimal clustering number (K value), and k=2 was finally selected as optimal clustering number. The sharp and clear boundaries showed stable and robust clustering for glioma patients (Figure 1C). To validate the two subclasses, we further performed individual PCA and t-SNE with decreased dimensions of features. We found that glioma patients were well distributed at two components (Figure 1D and Supplementary material Table S1, Cluster 1 and Cluster 2), and t-SNE also suggested that the samples presented two-dimensional distribution model (Figure 1E). The Kaplan-Meier survival curve indicated that the cluster 2 had worse OS than cluster 1 (Figure 1F). The clustering group was also associated with some clinical parameters (age, gender and grade and survival outcomes). HDAC1, HDAC4, HDAC5, HDAC6 and HDAC10 were highly expressed in cluster2, and HDAC1, HDAC2, HDAC3, and HDAC7, and HDAC9 were significantly highly expressed in cluster 1 (Figure 1G).

**Figure 1.**
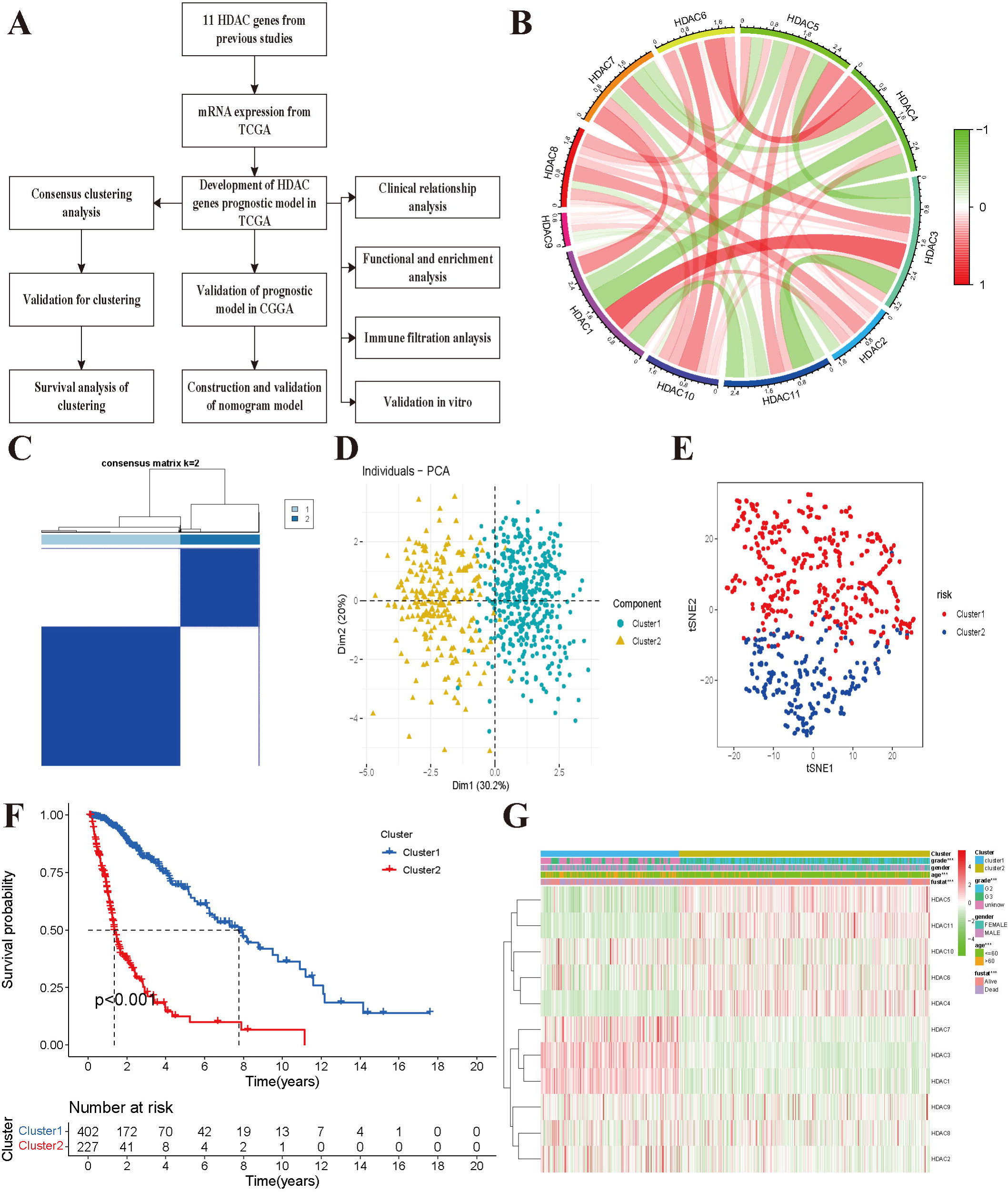
Glioma patients can be separated into two subclasses using HDAC genes. **A:** The flow chart of data analysis. **B:** The correlation circle plot among eleven HDAC genes. **C:** The consensus matrix plot identified the best grouping (k=2). **C**. Principal component analysis of glioma subclasses in the TCGA dataset. **D:** The corrected t-SNE2 analysis for two subclasses. **E:** The Kaplan-Meier survival curve for two subclasses in TCGA dataset.

### Development and validation of HDAC genes prognostic model in glioma

We first developed a prognostic model of OS in TCGA training dataset. The univariate cox regression indicated that high expressions of HDAC1, HDAC3, HDAC7 and HDAC9 were associated with poor OS in glioma, and the elevated expression of HDAC4, HDAC5, HDAC6 and HDAC11 were associated with favorable prognosis in glioma (Figure 2A). The HDAC2, HDAC2 and HDAC10 seemed not to be related to prognosis. Furthermore, the LASSO regression identified six HDAC genes (HDAC1, HDAC3, HDAC4, HDAC5, HDAC7 and HDAC9) that entered the final model (Figure 2B and 2C). Risk score of each sample was calculated according to the following formula: risk score=0.179*HDAC1_expression_ + 0.502*HDAC3_expression_ - 0.671*HDAC4_expression_ - 0.567*HDAC5_expression_ + 0.488*HDAC7_expression_ + 0.216*HDAC9_expression_ (Supplementary material Table S2). The glioma patients were categorized into high-risk group and low-risk group using the median of risk score (−0.360). The Kaplan-Meier curve showed that high-risk group had poorer OS than the low-risk group (P<0.001, Figure 2D). The risk score and survival time were also separately distributed (Figure 2E). PCA showed two obvious risk distribution patterns (Figure 2F).

**Figure 2.**
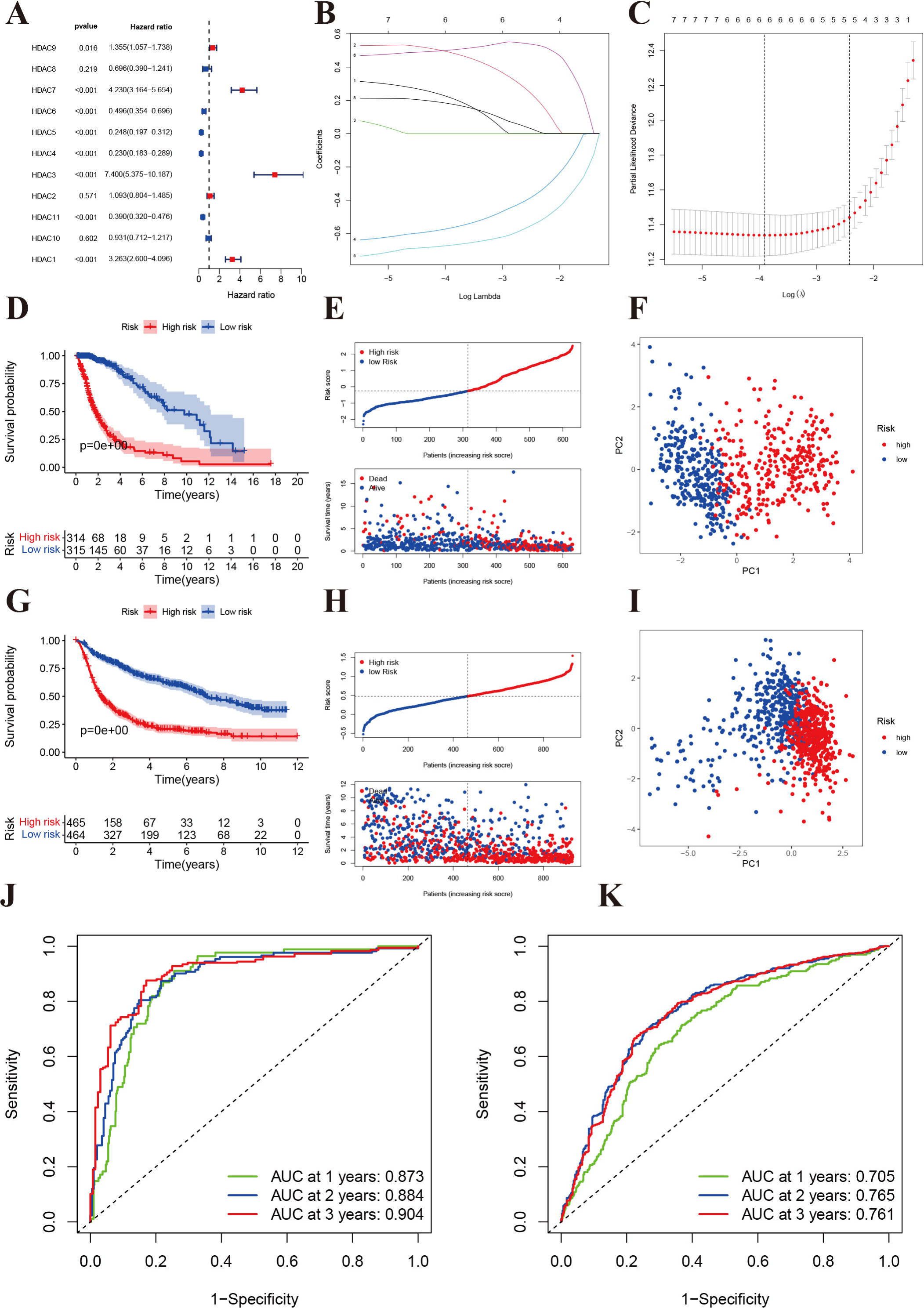
Development and validation of prognostic model based on HDAC genes. **A:** Forest plot of univariate cox regression for HDAC genes in glioma patients. **B:** LASSO regression of the 11 OS-related HDAC genes. **C:** Cross-validation for turning parameters selection in the LASSO regression. **D:** Kaplan-Meier survival curve of high- and low-risk groups from developed prognostic model based on 6 HDAC genes in TCGA. **E:** Distributions of risk scores and survival time of glioma patients in TCGA. **F:** PCA plot for high- and low-risk group in TCGA. **G:** Kaplan-Meier survival curve of high- and low-risk groups from validated prognostic model based on 6 HDAC genes in CGGA. **H:** Distributions of risk scores and survival time of glioma patients in CGGA. **I:** PCA plot for high- and low-risk group in CGGA. **J and K:** The receiver operating characteristic curve for predicting 1-year, 2-year, and 3-year survival rate of glioma patients in TCGA and CGGA.

Patients from CGGA dataset were used to validate the calculated risk score, and they were also separated into high- and low-risk groups according to the median calculated using the formula established in TCGA training set. Similarly, survival analysis suggested that high-risk groups had a poorer OS than low-risk group (P<0.001, Figure 2G), and risk score and survival time were also visually scattered (Figure 2H). Likewise, PCA showed two-dimensional distribution patterns (Figure 2I). The are under the curves (AUC) of 1-year, 2-year and 3-year were 0.873, 0.884 and .904 in the TCGA training set, respectively (Figure 2J). The AUCs of 1-year, 2-year and 3-year were 0.705, 0.765, and 0.761 in the CGGA validation set (Figure 2K).

### Stratified analysis

To further validate the prognostic model in different sub glioma patients, we performed stratified analysis in different sub-population. We found low-risk patients based on risk score had prolonged OS compared with high-risk group, which was not affected by different age, gender, histology, occurrence type, IDH codeletion status, 1p19q mutation, previous history of radiotherapy and chemotherapy (Figure 3A-3F, Figure 3H-3P). However, the OS showed insignificant differences for glioma patients with WHO II (Figure 3G), which means that the developed risk score may be inappropriate in such sub-population.

**Figure 3.**
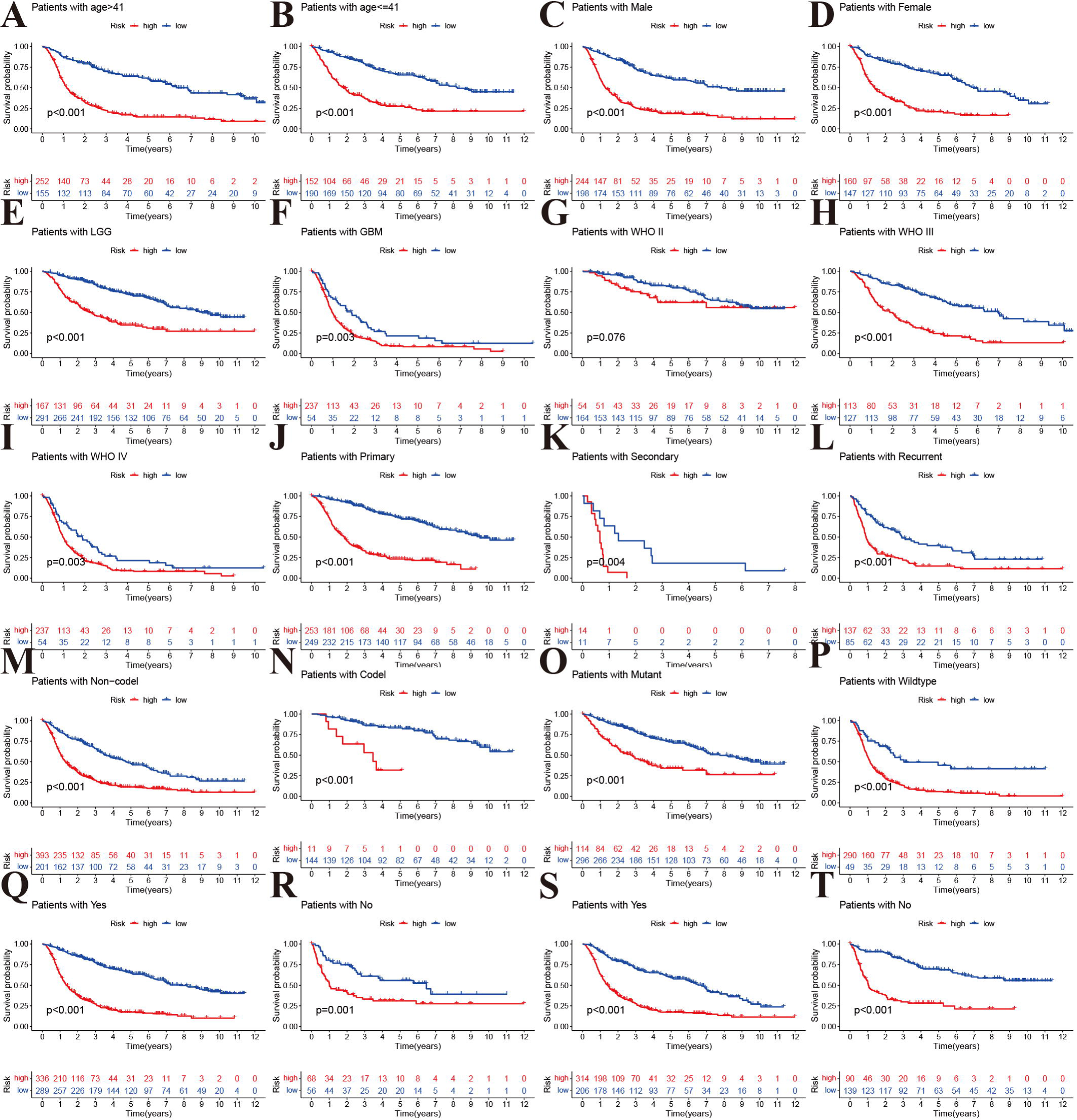
Stratified analyses of established HDAC-related genes prognostic model in TCGA. **A and B:** Age (>41 vs <=41). **C and D:** Gender (male vs female). **E and F:** Histology (LGG vs GBM). **G, H and I:** WHO stage (II, III and IV). **J-L:** Type of tumors (Primary, secondary vs recurrent). **M and N:** 1p19q (Non-codel and codel). **O and P:** mutant and wildtype. **Q and R:** Radiotherapy (Yes vs No). **S and T:** Chemotherapy (Yes vs No).

### Clinical correlation and independent and analysis

The analysis of expression difference indicated that all eleven HDAC genes showed significant differences between two subclasses (Figure 4A). The chi-square indicated that the high-risk patients tend to be GBM, WHO III/IV, older, occurrence or secondary, and have IDH mutation and 1p19q codeletion (*P*<0.05). No significant differences between high-and low-risk groups were observed for radiotherapy ratio and gender ratio (*P*>0.05, Figure 4B). We further compared the expression levels of six HDAC genes included in the prognostic model, and found HDAC1 (*P*<0.001), HDAC3 (*P*<0.001), HDAC7(*P*<0.001) and HDAC9 were highly expressed in high-risk group, while HDAC4 (*P*<0.001) and HDAC5 (*P*<0.001) were lowly expressed in high-risk group (*P*<0.001). Then we compared the risk score differences among different clinical parameters. Our results indicated that patients with >41 years old, advanced WHO stage and grade had higher risk scores (*P*<0.005, Figure 4C-4F). Patients with IDH mutation and 1p19q codeletion had lower risk score (*P*<0.05, Figure 4H and 4I). However, the risk score showed no significant differences among different gender (Figure 4D), recurrent or secondary (Figure 4G), or radiotherapy status (Figure 4J). Patients who received chemotherapy also have higher risk score compared those without chemotherapy (Figure 4K).

**Figure 4.**
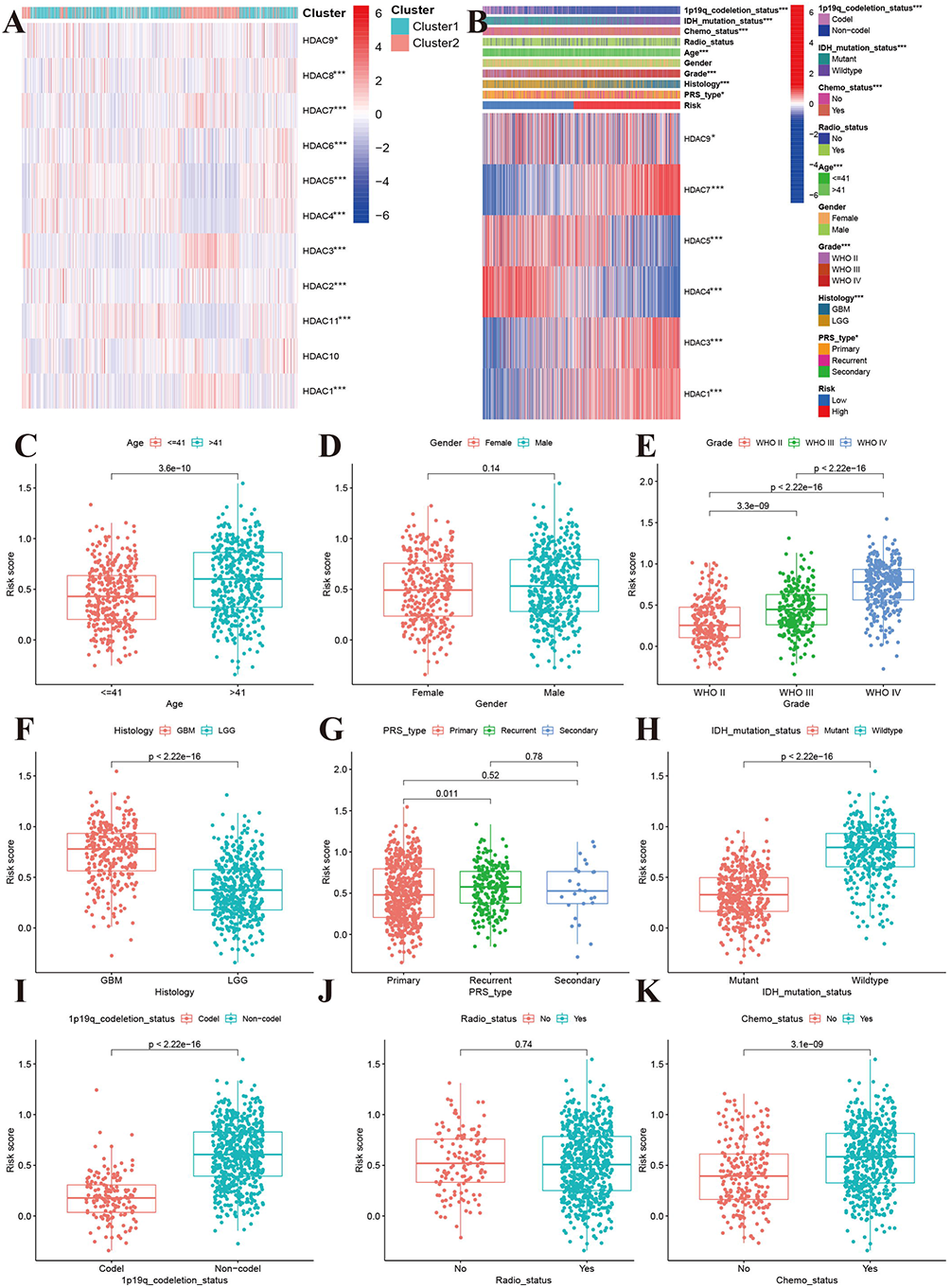
Association between HDAC genes and clinical characteristics in glioma patients. **A:** Heatmap indicated the expression of HDAC genes between two subclasses. **B:** Heatmap of associations among risk stratifications and clinical parameters and six HDAC gens expression. Comparisons of risk score among different clinical parameters: **C:** age (>41 vs <=41), **D:** gender (male vs female), **E:** WHO stage (II, III, IV). **F:** histology (LGG vs GBM). **G:** PRS type (primary, recurrent, and secondary). **H:** IDH mutation status (mutant vs wild type). **I:** 1p19q codeletion status (codel vs non-codel). **J:** radiotherapy status (No vs Yes). **K:** chemotherapy (No vs Yes).

To investigate whether risk score was an independent prognostic factor for glioma patients or not, we performed univariate and multiple variates cox regression in the TCGA training set and CGGA validation set. In TCGA dataset, the univariate and multiple variate cox regression indicated that risk score was associated with OS in glioma patients (univariate: HR=2.084, 95%CI: 1.890-2.297, P<0.001, Figure 5A; multiple: HR=1.425, 95%CI: 1.247-1.629, P<0.001, Figure 5B). The ROC result showed the risk score have optimal predictive ability (AUC=0.828) for 5-year OS (Figure 5C). Similarly, the elevated risk score also increased the risk of poor OS (univariate: HR=7.801 95%CI: 5.887-10.338, P<0.001, Figure 5D; multiple: HR=2.184, 95%CI: 1.484-3.213, P<0.001, Figure 5E). The AUC was 0.808 that was higher than any of other clinical parameters (Figure 5F). Besides, recurrence, advanced grade and age, were also risk factor for poor OS, while patients who received chemotherapy and have IDH mutation and 1p19q codeletion had favorable OS (Figure 5E).

**Figure 5.**
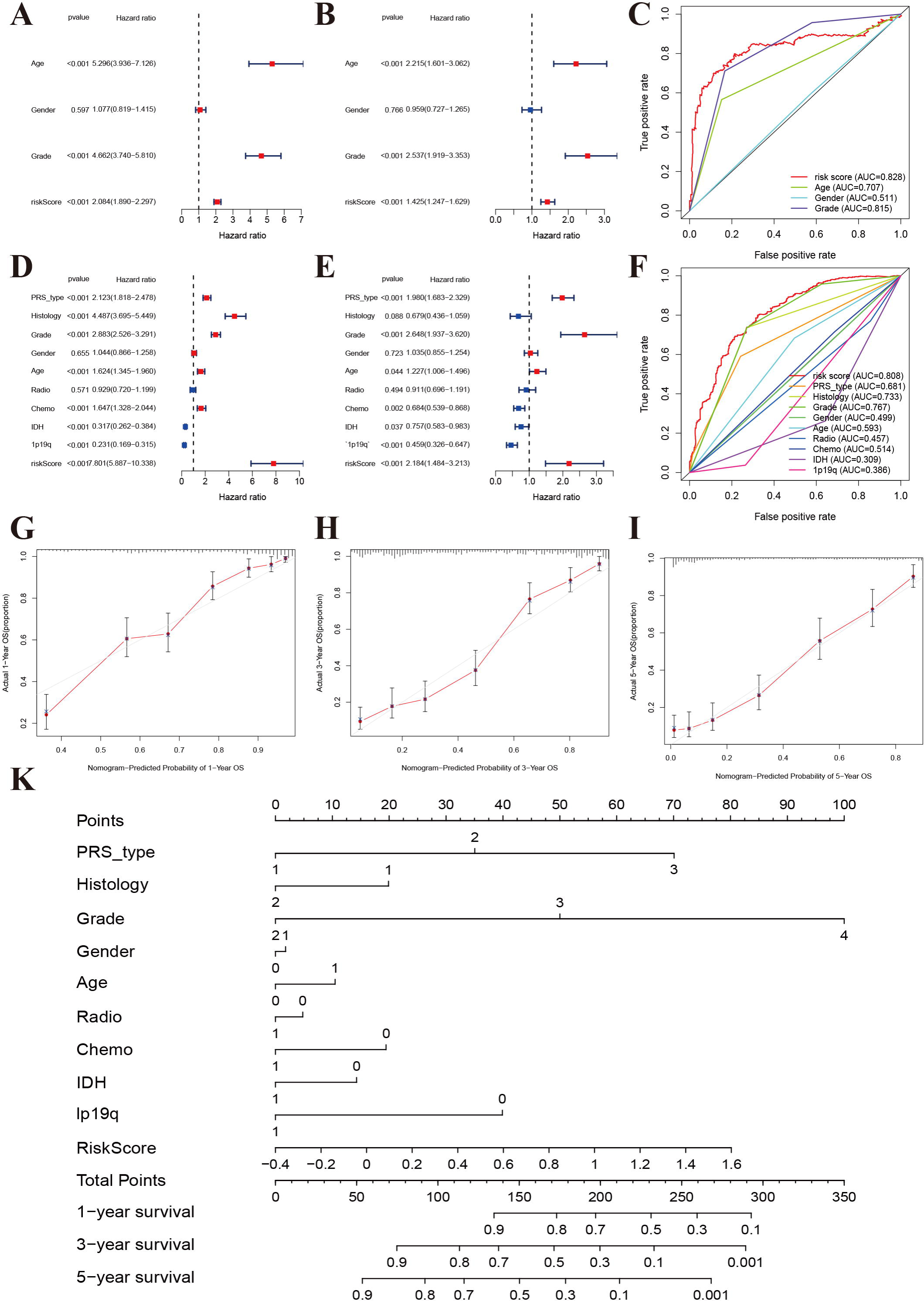
Independent prognosis analyses of HDAC-related genes model. **A and B:** univariate and multivariate cox regression of risk score based on HDAC genes in TCGA. **C:** The receiver operating characteristic curve of risk score for predicting 5-year survival rate in TCGA. **D and E:** univariate and multivariate cox regression of risk score based on HDAC genes in CGGA. **F:** The receiver operating characteristic curve of risk score for predicting 5-year survival rate in CGGA. **G-I:** Calibration curves of 1-eyar, 3-year, and 5-year OS in TCGA. **K:** Nomograph model established in CGGA cohort.

### Functional, pathway enrichment and mutations analysis

To explore the functional and pathway enrichment of high-and low-risk groups, we performed GO and KEGG analysis. We first identified potential differently expressed gens between high- and low-risk groups (Log fold change >1, *P*<0.05). We finally identified 2598 differently expressed genes, including 1723 upregulated genes and 875 downregulated genes in the high-risk groups (Supplementary material Table S3). GO enrichment analysis indicated that high risk group was mainly enriched in immune-related function in biological process, collagen and lumen in cellular component,, and antigen binding, extracellular matrix structural constituent, some regulator, receptors and inhibitors binding in molecular function (Figure 6A). The KEGG pathways analysis showed that the high-risk group was involved PI3K-Akt signaling pathways, AGE-RAGE signaling, HIF-1 signaling, Relaxin signaling, and p53 signaling. Focal adhesion, ECM-receptor and cytokine-cytokine receptor interaction, cell cycle and pyrimidine metabolism were also significantly enriched (Figure 6B). The occurrence of glioma was involved in the integrations of multiple molecular functions and signaling pathways.

**Figure 6.**
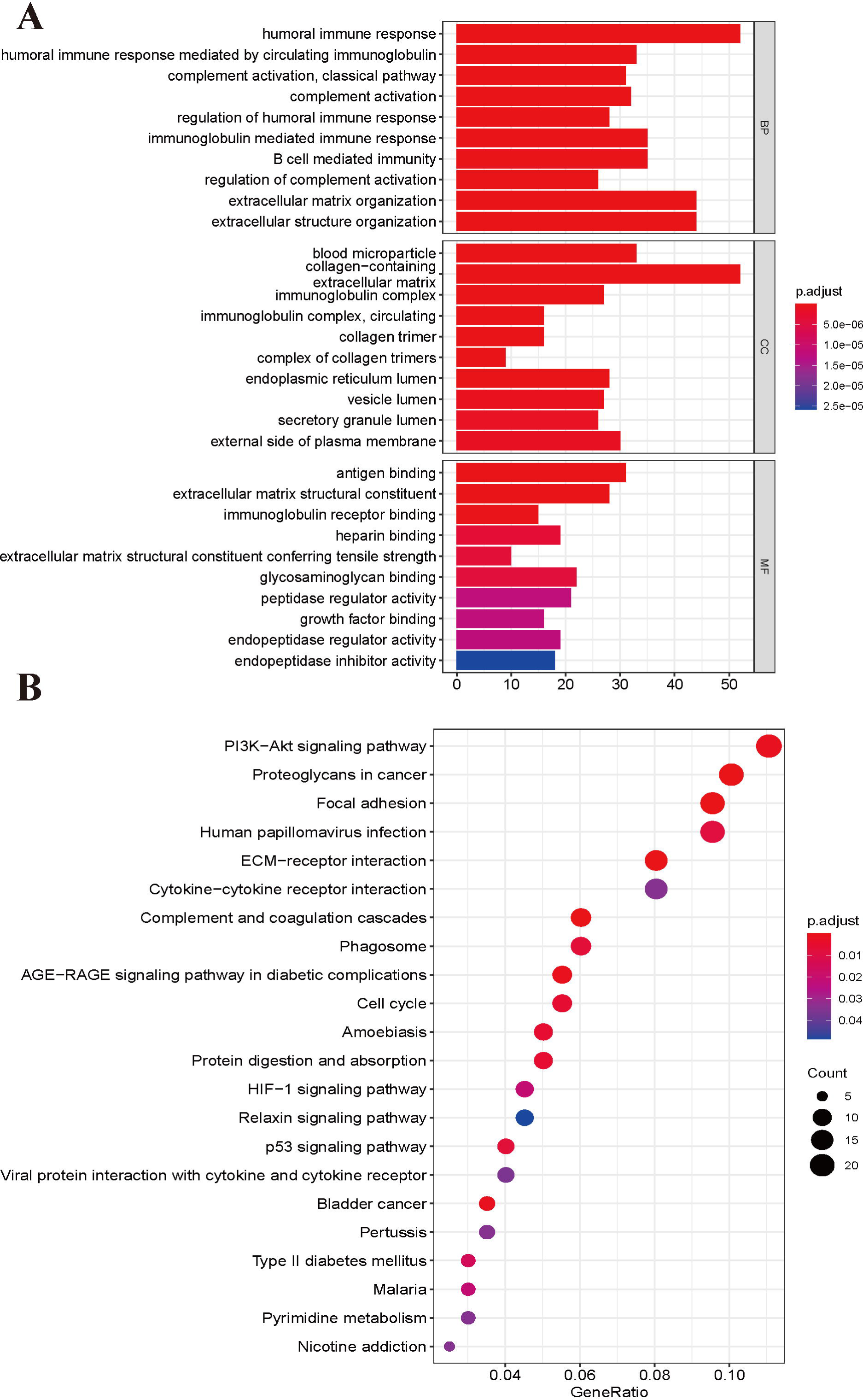
GO enrichment (**A**) and KEGG pathways analysis (**B**) based on differently expressed genes between high- and low risk groups.

We also explored the gene mutation differences between high-risk and low-risk groups. Our results indicated that there are significant differences in gene mutation between high- and low-risk groups. The high-risk group showed high gene alteration in EGFR, PTEN, FLG, PKHD1(Figure 7A), while the gene alteration rate of IDH, ATRX, CIC, was higher in the low-risk group than in the high-risk group (Figure 7B). High- and low-risk groups showed similar results in variant classification, variant type, SNV class (Figure 7C and 7D). Furthermore, HYDIN-PI3CA, AHNAK2-SPTA1, COL6A3-PTEN, and IDH1-TP53 showed highly co-occurrence, and PTEN-TTN, IDH1-EGFR showed mutually exclusive in high-risk groups. The SSPO-HMCN1, LRP2, NIPBL, MYH1-CIC, TTN, MUC16, APOB, RYR2, DNMT3A, NIPBL-IDH2, APOB, NOTCH1, LRP2 showed highly co-occurrence (Figure 7E). The IDH2-TP53, PI3CA-TP53, CIC-TP53 showed mutually exclusive in low-risk group (Figure 7F).

**Figure 7.**
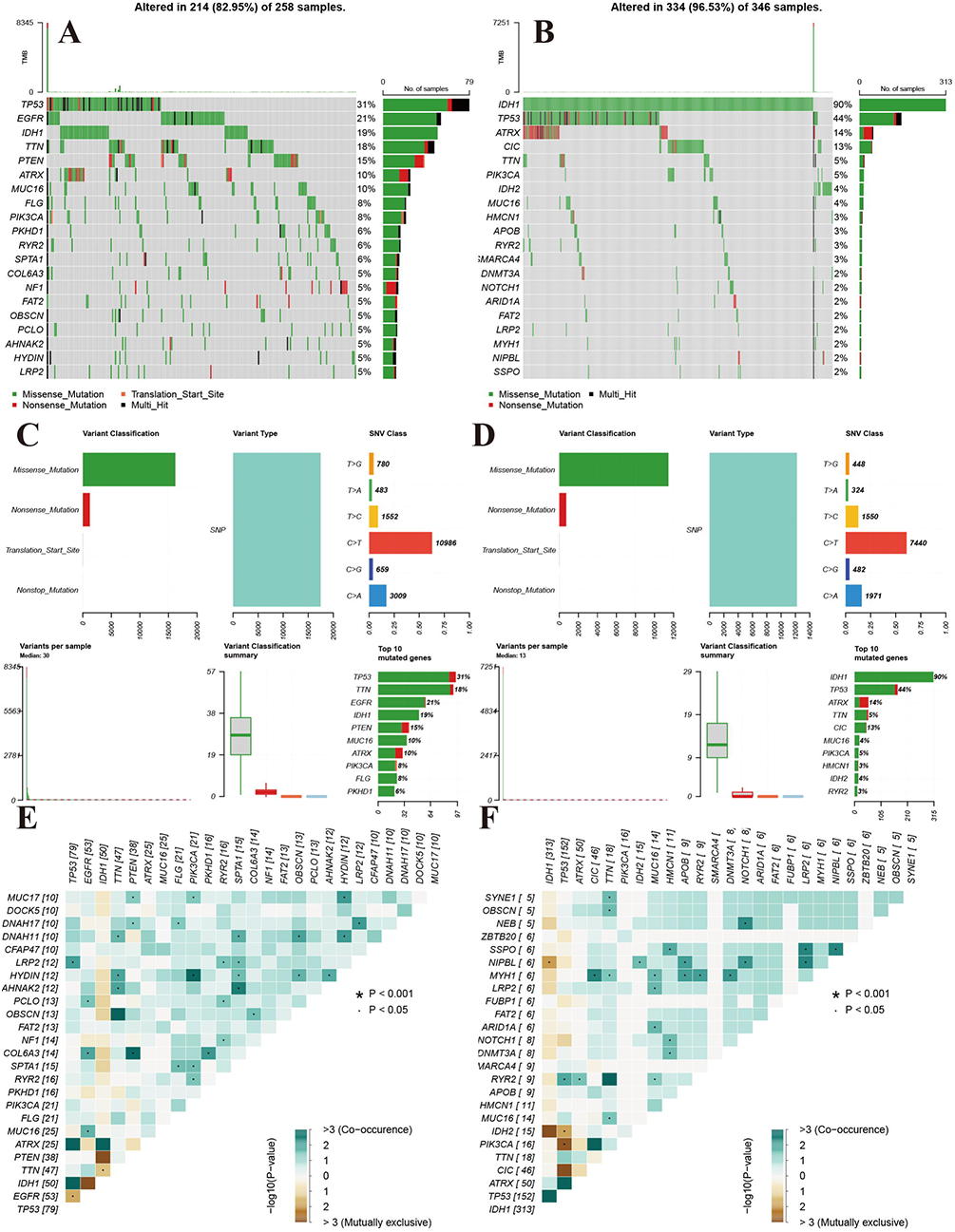
Landscape of mutation profiles between high- and low-risk groups. **A and B:** Waterfall plots of mutation information in each sample. **C and D:** Bar graph of variant classification. **E and F:** somatic interactions plot (co-occurrence and exclusive)

### Immune filtration, tumor microenvironment, and drug sensitivity analysis

To explore the immune response difference between high- and low-risk group, we compared the immune filtration cells and immune-related pathways between high- and low-risk groups. Our results indicated that the aDCs, B cells, CD8+ Tcells, iDCs, macrophages, NK cells, pDCS, T helper cells, Tfh, Th1 cell, Th2 cells, TIL, and Treg were highly expressed in high-risk group, and no significant differences were observed in DCs, Mast cells and neutrophils between different risk groups (Figure 8A). All immune-related pathways were highly enriched in high-risk group (Figure 8B). we also found that the ESTIMATE score, immune score, and stromal score were higher in the high-risk group than in the low-risk group (Figure 8C-8E). The Pearson correlation analysis indicated that macrophages M0, M1, and M2 showed positive associations with risk score (Figure 8F, 8I, and 8J), while monocytes, NK cells activated, mast cells activated showed negative associations with risk score (Figure 8G, 8H, 8K).

**Figure 8.**
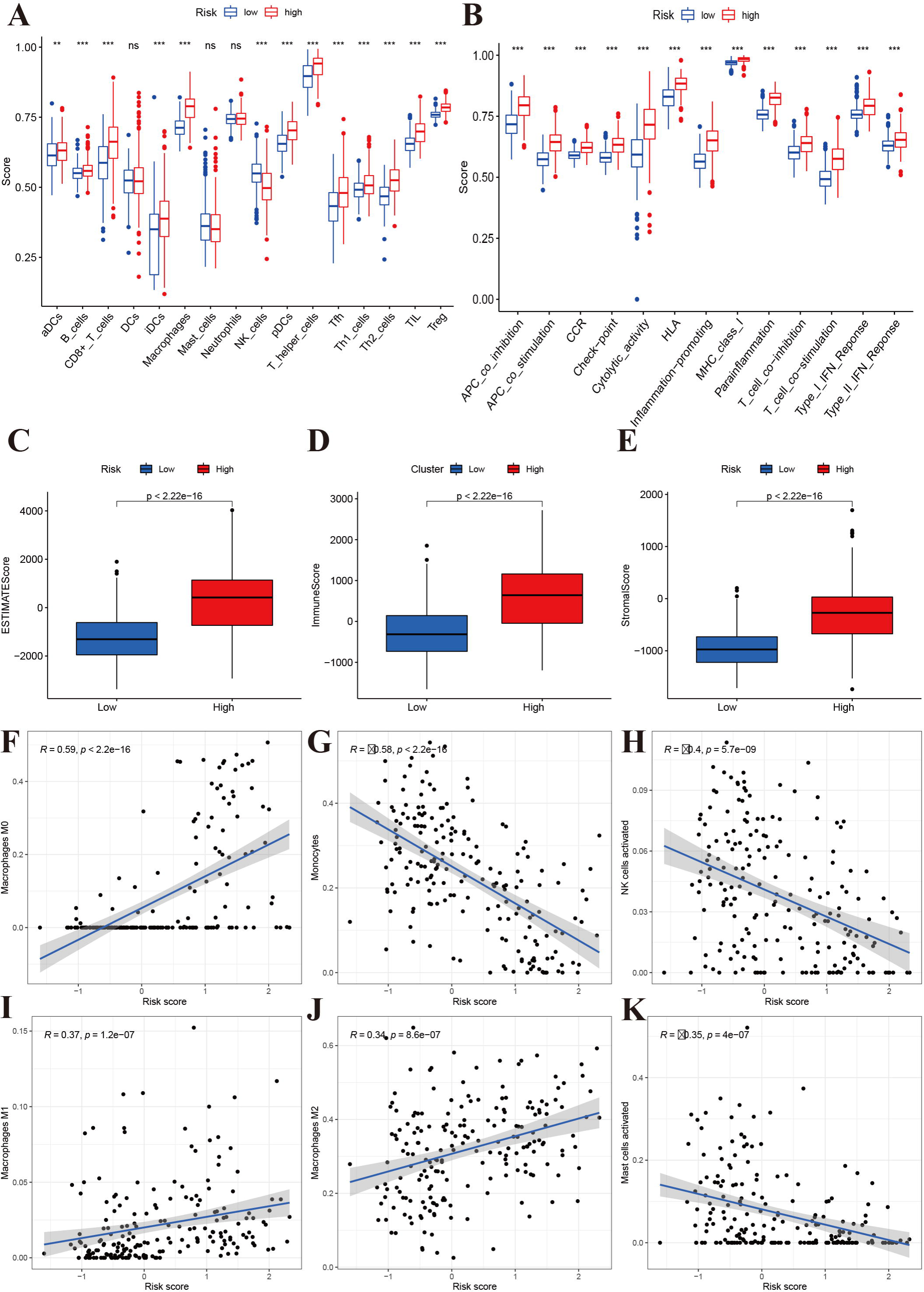
Immune status analysis between high- and low-risk group. **A:** The ssGSEA scores of immune cells. **B:** The ssGSEA scores of immune-related functions. **C-E:** Comparisons of Estimated, immune and stromal score between high-and low-risk group. **F-K:** Correlation between risk score and immune markers (Macrophages M0, Monocytes, NK cells activated, Macrophages M1, M2, and Mast cells activated) in glioma patients.

To explore the potential small molecular drug related with HDAC genes, we performed Pearson correlation analysis between HDAC genes expression and some small molecular drug (Figure 9). Our results indicated that HDAC7 showed significant drug resistance with Selumetinib, Cobimetinib, Trametinib, PD-98059, Dabrafenib, and Dolastatin 10, while Everolimus, Rapamycin, and Temsirolimus showed positive associations with HDAC7. PX-316, Chelerythrine, Selumetinib were positively associated with HDAC4 expression. HDAC9 showed a negative correlation with By-product o. Chelerythrine and Acrichine were also positively associated with HDAC1. These results provided some potential clues for HDAC-targeted treatment in glioma.

**Figure 9.**
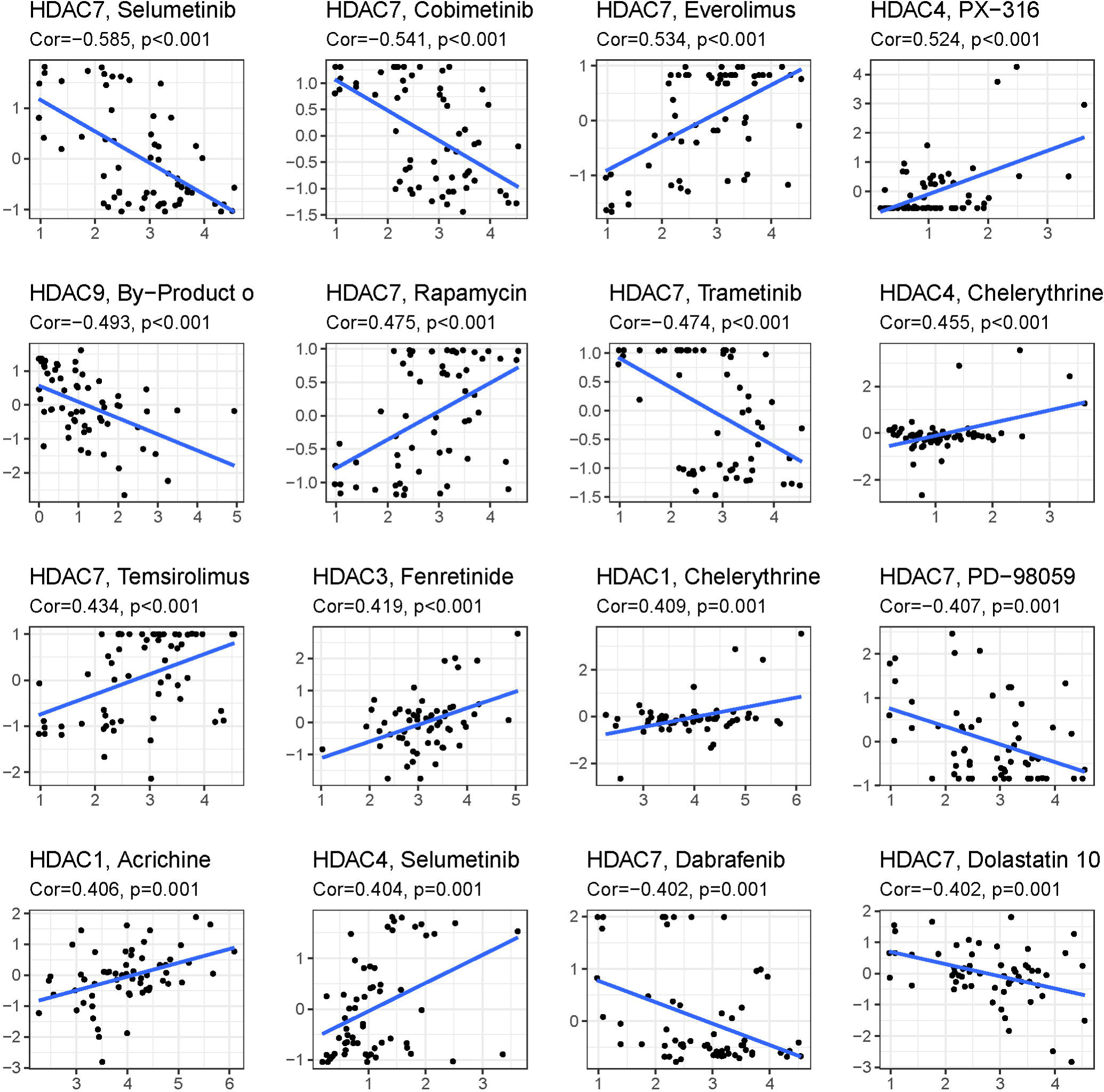
Top 16 kinds of drug associated with HDAC member.

### Validation of HDAC genes in glioma and non-tumor tissue

The qPCR was adopted methods to detect the expression of HDAC1, HDAC3, HDAC4, HDAC5, HDAC7 and HDCA9 of non-tumor and glioma tissue. The results were presented in Figure 10. The expression levels of HDAC1 (Figure 10A), HDAC3 (Figure 10B), HDAC7 (Figure 10E), and HDAC9 (Figure 10F) was significantly elevated in the glioma patients compared with non-tumor group. However, the expression levels of HDAC4 and HDAC5 were lower in the glioma patients than in the non-tumor control groups (Figure 10 C and 10D). This result was consistent with the role of these HDAC genes in prognosis.

**Figure 10.**
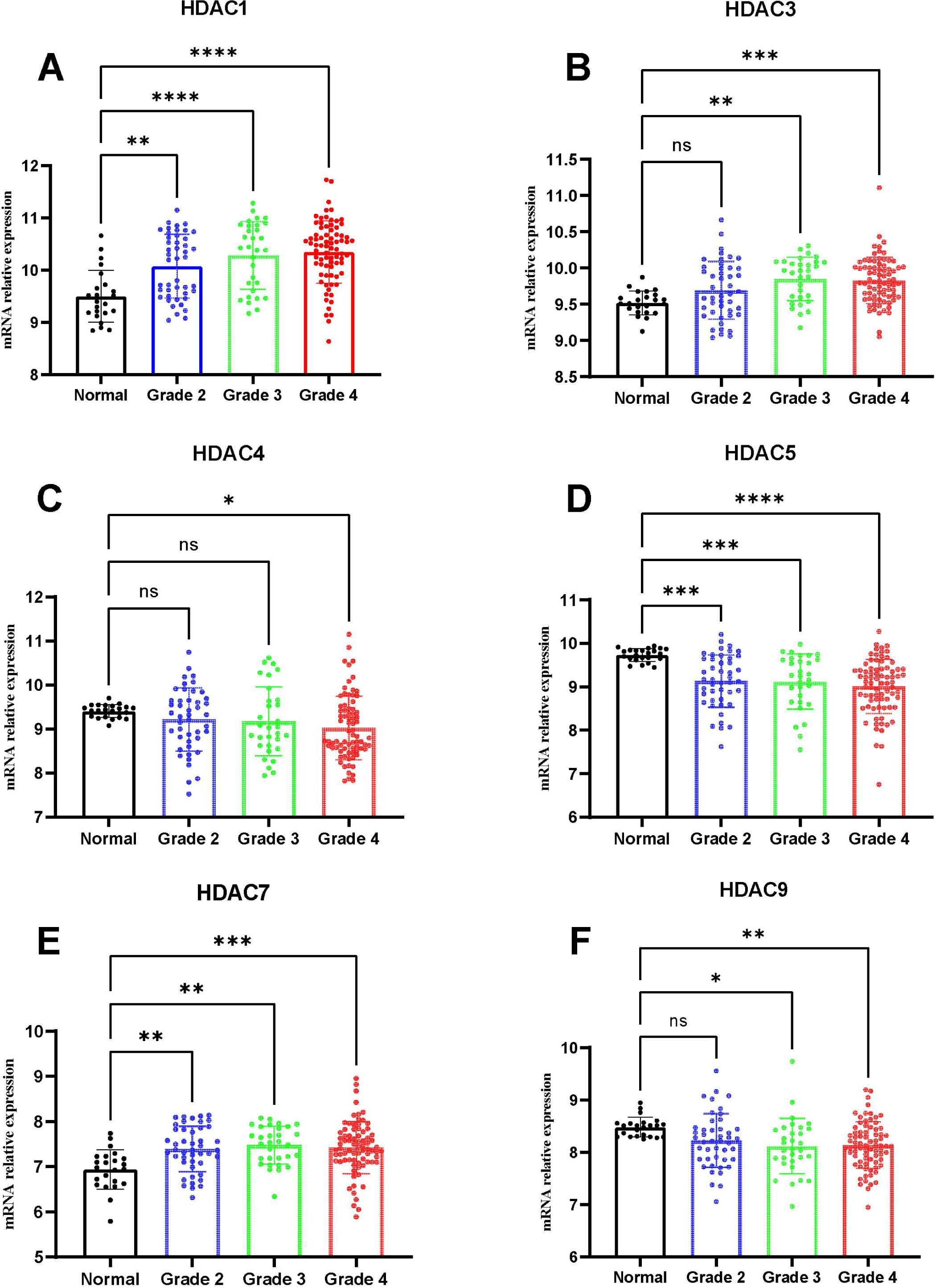
Expression of HDAC genes in glioma and non-tumor tissue. **A:** HDAC1, **B:** HDAC3, **C:** HDAC4, **D:** HDAC5, **E:** HDAC7, **F:** HDAC9

## Discussion

HDACs, as key enzymes that catalyze the acetylation of histones, are involved in many processes such as the growth and proliferation of malignant tumor cells, expression regulation, etc[16]. In the epigenetic research of tumor occurrence and development, it has gradually attracted academic wide attention in the world. Most of the current research focuses on the chemical modification and structural modification of the existing antitumor drugs with HDACs inhibitory activity to enhance the therapeutic effect of the drug and alleviate the toxic and side effects[17]. There are currently few antitumor drugs designed to act on specific targets and specific pathways. At the same time, it is of great of need to explore the molecular signatures for more understanding of biological relationship between tumor genotype and phenotypes.

In the present study, we found that glioma patients can be divided into two subclasses based on eleven HDAC genes, and patients from two subclasses had markedly different survival outcomes. Then, using six HDAC genes (HDAC1, HDAC3, HDAC4, HDAC5, HDAC7, and HDAC9), we established a prognostic model in glioma patients, and this prognostic model was well validated in an independent cohort population. Furthermore, the calculated risk score from six HDACA genes expression was suggested to be an independent prognostic factor, which can predict the five-year overall survival of glioma patients well. High-risk patients can be attributed to multiple complex function and molecular signaling pathways, and the genes alterations of high- and low-risk patients were significantly different. We also found that different survival outcomes of high- and low-risk patients could be involved in the differences of immune filtration level and tumor microenvironment. Subsequently, we identified several small molecular compounds that could be favorable for glioma patients’ treatment. And finally, we validated the expression levels of HDAC genes from prognostic model using glioma and non-tumor tissues samples. Our study provided new and simple molecular subtypes and prognosis prediction methods, and added more understanding for biology and molecular mechanisms of glioma.

We identified six HDAC genes in established prognostic model. HDAC1 and HDAC3 belongs to Class I of HDAC. Previous studies had indicated that HDAC1 was over-expressed in diverse human malignancies, such as prostate cancer, breast cancer, liver cancer, and lung cancer[18-21]. HDAC1 was also highly expressed in glioma tissue, and highly expression of glioma is associated with glioma cell proliferation, migration, invasion, angiogenesis, and poor prognosis[22]. Besides, it was suggested that increased activation of HDAC1/2/6 and Sp1 underlies therapeutic resistance and tumor growth in glioblastoma[23]. We also found expression of HDAC1 was elevated in glioma tissues and be associated with poor prognosis. HDAC3 has become a focus in recent research, many scholars worldwide have found that it plays a role of carcinogenesis in human tumors. After the expression of HDAC3 is reduced by the inhibitors, the growth and invasive ability of human glioma cell appears significantly weakened, which provides a new way for the cancer treatment[24]. HDAC4, HDAC5, HDAC7, and HDAC9 belong to Class II of HDACs[25]. HDAC4 is frequently dysregulated in human malignancies, and we also confirmed the downregulated expression in glioma tissues. However, previous studies reported that HDAC4 was significantly upregulated in glioma tissues. The proliferation, adenosine triphosphate (ATP) levels and invasion ability were substantially enhanced in U251 cells with HDAC4 overexpression, and suppressed in U251 cells with a knockdown of HDAC4 compared with that in U251 cells transfected with the negative control[26, 27]. This may be associated with glioma grade, stage and histology, and further research was required. Just like HDAC4, HDAC5 was also found to be lowly expressed in glioma tissue. HDAC7 plays an oncogene role in glioma. It was reported that ZNF326 could activate HDAC7 transcription by binding to a specific promoter region via its transcriptional activation domain and zinc-finger structures in glioma cell[28]. Furthermore, ZNF326 was not only highly expressed in glioma but was also positively correlated with the expression of HDAC7, which identified the oncogene role of HDAC7[29]. HDAC9, like most class II HDACs, has a conserved histone deacetylase domain, catalyzes the removal of acetyl moieties in the N-terminal tail of histones, and possesses a long regulatory N-terminal domain to interact with tissue-specific transcription factors and co-repressors, the amino-terminal domain contains highly conserved serine residues that are subjected to phosphorylation[30]. Signal-dependent phosphorylation of HDAC9 is a critical event that determines whether it is localized in the cytoplasm or nucleus. High expression of HDAC9 has been reported in many cancers[31-33]. In glioma, the high expression of HDAC9 can promote the proliferation and tumor formation, and accelerated cell cycle in part by potentiating the EGFR signaling pathways[34]. With the emerging and rapid development of disciplines such as structural biomechanics and computer-aided drug design, the development of new HDACs inhibitors with antitumor activity targeting HDACs is bound to have a very broad development space and development prospects.

## Conclusions

Sum it up, our results reveled the clinical utility and potential molecular mechanisms of HDAC genes in glioma. Model based on six HDAC genes (HDAC1, HDAC3, HDAC4, HDAC5, HDAC7, and HDAC9) can predict the overall survival of glioma patients well, which can be served as potential therapeutic targets. The future research should validate this model in a large cohort population, and experiments in vivo and vitro will favor the understanding of molecular mechanisms of glioma.

## Data Availability

The TCGA data can be obtained from the https://portal.gdc.cancer.gov/, and CGGA data can be available from the Chinese Glioma Genome Atlas (http://www.cgga.org.cn/). The expression levels of HDAC genes from non-tumor and glioma patients can be available from the GEO (GSE4290).

## Competing interests

The authors declare that they have no conflict of interest.

## Funding

This study was supported by the National Natural Science Foundation of China (LZZ: 82003239), and Science Foundation of Xiangya Hospital for Young Scholar (LZZ: 2018Q012), and National Natural Science Foundation of China (SLF:81974466).

## Authors’ contributions

ZZL designed this study and directed the research group in all aspects, including planning, execution, and analysis of the study. LAB drafted the manuscript. LYY, NL, SL, LAB collected the data. LZZ provided the statistical software, performed the data analysis, SL arranged the Figures and Tables. SLF revised the manuscript. All authors have read and approved the final version of the manuscript.

## Supplementary materials

**Table S1** Clustering of glioma based on 11 HDAC genes in TCGA

**Table S2** Coefficient of HDAC in the included model

**Table S3** Differential expressed genes between high-and low-risk group

## Notes

### Competing Interest Statement

The authors have declared no competing interest.

## References

1. Weller M, Wick W, Aldape K, Brada M, Berger M, Pfister SM, et al. (2015) Glioma. Nat Rev Dis Primers 1, 15017, https:10.1038/nrdp.2015.17

2. Chen R, Smith-Cohn M, Cohen AL, Colman H (2017) Glioma Subclassifications and Their Clinical Significance. Neurotherapeutics 14, 284–297, https:10.1007/s13311-017-0519-x

3. Kan LK, Drummond K, Hunn M, Williams D, O’Brien TJ, Monif M (2020) Potential biomarkers and challenges in glioma diagnosis, therapy and prognosis. BMJ Neurol Open 2, e69, https:10.1136/bmjno-2020-000069

4. Bai J, Varghese J, Jain R (2020) Adult Glioma WHO Classification Update, Genomics, and Imaging: What the Radiologists Need to Know. Top Magn Reson Imaging 29, 71–82, https:10.1097/RMR.0000000000000234

5. Snape TJ, Warr T (2015) Approaches toward improving the prognosis of pediatric patients with glioma: pursuing mutant drug targets with emerging small molecules. Semin Pediatr Neurol 22, 28–34, https:10.1016/j.spen.2014.12.003

6. Hanahan D, Weinberg RA (2011) Hallmarks of cancer: the next generation. Cell 144, 646–674, https:10.1016/j.cell.2011.02.013

7. Dawson MA, Kouzarides T (2012) Cancer epigenetics: from mechanism to therapy. Cell 150, 12–27, https:10.1016/j.cell.2012.06.013

8. Nebbioso A, Tambaro FP, Dell’Aversana C, Altucci L (2018) Cancer epigenetics: Moving forward. Plos Genet 14, e1007362, https:10.1371/journal.pgen.1007362

9. Hai R, He L, Shu G, Yin G (2021) Characterization of Histone Deacetylase Mechanisms in Cancer Development. Front Oncol 11, 700947, https:10.3389/fonc.2021.700947

10. Mahmud I, Liao D (2015) Microarray gene expression profiling reveals potential mechanisms of tumor suppression by the class I HDAC-selective benzoylhydrazide inhibitors. Genom Data 5, 257–259, https:10.1016/j.gdata.2015.06.019

11. Li S, Paterno GD, Gillespie LL (2013) Nuclear localization of the transcriptional regulator MIER1alpha requires interaction with HDAC1/2 in breast cancer cells. Plos One 8, e84046, https:10.1371/journal.pone.0084046

12. Thangaraju M, Carswell KN, Prasad PD, Ganapathy V (2009) Colon cancer cells maintain low levels of pyruvate to avoid cell death caused by inhibition of HDAC1/HDAC3. Biochem J 417, 379–389, https:10.1042/BJ20081132

13. Ropero S, Esteller M (2007) The role of histone deacetylases (HDACs) in human cancer. Mol Oncol 1, 19–25, https:10.1016/j.molonc.2007.01.001

14. Cheng F, Zheng B, Wang J, Zhao G, Yao Z, Niu Z, et al. (2021) Comprehensive analysis of a new prognosis signature based on histone deacetylases in clear cell renal cell carcinoma. Cancer Med 10, 6503–6514, https:10.1002/cam4.4156

15. Sun L, Hui AM, Su Q, Vortmeyer A, Kotliarov Y, Pastorino S, et al. (2006) Neuronal and glioma-derived stem cell factor induces angiogenesis within the brain. Cancer Cell 9, 287–300, https:10.1016/j.ccr.2006.03.003

16. Rroji O, Kumar A, Karuppagounder SS, Ratan RR (2021) Epigenetic regulators of neuronal ferroptosis identify novel therapeutics for neurological diseases: HDACs, transglutaminases, and HIF prolyl hydroxylases. Neurobiol Dis 147, 105145, https:10.1016/j.nbd.2020.105145

17. Chen X, He Y, Fu W, Sahebkar A, Tan Y, Xu S, et al. (2020) Histone Deacetylases (HDACs) and Atherosclerosis: A Mechanistic and Pharmacological Review. Front Cell Dev Biol 8, 581015, https:10.3389/fcell.2020.581015

18. Burdelski C, Ruge OM, Melling N, Koop C, Simon R, Steurer S, et al. (2015) HDAC1 overexpression independently predicts biochemical recurrence and is associated with rapid tumor cell proliferation and genomic instability in prostate cancer. Exp Mol Pathol 98, 419–426, https:10.1016/j.yexmp.2015.03.024

19. Zhang Y, Nalawansha DA, Herath KE, Andrade R, Pflum M (2021) Differential profiles of HDAC1 substrates and associated proteins in breast cancer cells revealed by trapping. Mol Omics 17, 544–553, https:10.1039/d0mo00047g

20. Rivas M, Johnston MN, Gulati R, Kumbaji M, Margues AT, Timchenko L, et al. (2021) HDAC1-Dependent Repression of Markers of Hepatocytes and P21 Is Involved in Development of Pediatric Liver Cancer. Cell Mol Gastroenterol Hepatol 12, 1669–1682, https:10.1016/j.jcmgh.2021.06.026

21. Dong ZY, Zhou YR, Wang LX (2018) HDAC1 is indirectly involved in the epigenetic regulation of p38 MAPK that drive the lung cancer progression. Eur Rev Med Pharmacol Sci 22, 5980–5986, https:10.26355/eurrev_201809_15932

22. Fan Y, Peng X, Wang Y, Li B, Zhao G (2021) Comprehensive Analysis of HDAC Family Identifies HDAC1 as a Prognostic and Immune Infiltration Indicator and HDAC1-Related Signature for Prognosis in Glioma. Front Mol Biosci 8, 720020, https:10.3389/fmolb.2021.720020

23. Yang WB, Hsu CC, Hsu TI, Liou JP, Chang KY, Chen PY, et al. (2020) Increased activation of HDAC1/2/6 and Sp1 underlies therapeutic resistance and tumor growth in glioblastoma. Neuro Oncol 22, 1439–1451, https:10.1093/neuonc/noaa103

24. Zhong S, Fan Y, Wu B, Wang Y, Jiang S, Ge J, et al. (2018) HDAC3 Expression Correlates with the Prognosis and Grade of Patients with Glioma: A Diversification Analysis Based on Transcriptome and Clinical Evidence. World Neurosurg 119, e145–e158, https:10.1016/j.wneu.2018.07.076

25. Hou F, Li D, Yu H, Kong Q (2018) The mechanism and potential targets of class II HDACs in angiogenesis. J Cell Biochem 119, 2999–3006, https:10.1002/jcb.26476

26. Cai JY, Xu TT, Wang Y, Chang JJ, Li J, Chen XY, et al. (2018) Histone deacetylase HDAC4 promotes the proliferation and invasion of glioma cells. Int J Oncol 53, 2758–2768, https:10.3892/ijo.2018.4564

27. Wang Y, Xia Y, Hu K, Zeng M, Zhi C, Lai M, et al. (2019) MKK7 transcription positively or negatively regulated by SP1 and KLF5 depends on HDAC4 activity in glioma. Int J Cancer 145, 2496–2508, https:10.1002/ijc.32321

28. Yu X, Wang M, Wu J, Han Q, Zhang X (2019) ZNF326 promotes malignant phenotype of glioma by up-regulating HDAC7 expression and activating Wnt pathway. J Exp Clin Cancer Res 38, 40, https:10.1186/s13046-019-1031-4

29. Wang M, Han Q, Su Z, Yu X (2020) Transcription Factor ZNF326 Upregulates the Expression of ERCC1 and HDAC7 and its Clinicopathologic Significance in Glioma. Lab Med 51, 377–384, https:10.1093/labmed/lmz075

30. Fernandez-Ruiz I (2020) HDAC9 linked to aortic calcification. Nat Rev Cardiol 17, 6–7, https:10.1038/s41569-019-0308-9

31. Xu G, Li N, Zhang Y, Zhang J, Xu R, Wu Y (2019) MicroRNA-383-5p inhibits the progression of gastric carcinoma via targeting HDAC9 expression. Braz J Med Biol Res 52, e8341, https:10.1590/1414-431X20198341

32. Linares A, Assou S, Lapierre M, Thouennon E, Duraffourd C, Fromaget C, et al. (2019) Increased expression of the HDAC9 gene is associated with antiestrogen resistance of breast cancers. Mol Oncol 13, 1534–1547, https:10.1002/1878-0261.12505

33. Liang Z, Feng A, Shim H (2020) MicroRNA-30c-regulated HDAC9 mediates chemoresistance of breast cancer. Cancer Chemother Pharmacol 85, 413–423, https:10.1007/s00280-019-04024-9

34. Zhang Y, Wu D, Xia F, Xian H, Zhu X, Cui H, et al. (2016) Downregulation of HDAC9 inhibits cell proliferation and tumor formation by inducing cell cycle arrest in retinoblastoma. Biochem Biophys Res Commun 473, 600–606, https:10.1016/j.bbrc.2016.03.129

